# COVID-19 Bimodal Clinical and Pathological Phenotypes

**DOI:** 10.1101/2021.09.03.21262841

**Authors:** Sabrina S Batah, Maíra N Benatti, Li Siyuan, Wagner M Telini, Jamile Barbosa, Marcelo B Menezes, Tales R Nadai, Keyla S G Sá, Chirag M. Vaswani, Sahil Gupta, Dario S Zamboni, Danilo T Wada, Rodrigo T Calado, Renê D R Oliveira, Paulo Louzada-Junior, Maria Auxiliadora-Martins, Flávio P Veras, Larissa D Cunha, Thiago M Cunha, Rodrigo Luppino-Assad, Marcelo L Balancin, Sirlei S Morais, Ronaldo B Martins, Eurico Arruda, Fernando Chahud, Marcel Koenigkam-Santos, Andrea A Cetlin, Fernando Q Cunha, Claudia dos Santos, Vera L Capelozzi, Junya Fukuoka, Rosane Duarte-Achcar, Alexandre T Fabro

## Abstract

**Background:** Patients with coronavirus disease-2019 (COVID-19) present varying clinical complications. Different viral load and host response related to genetic and immune background are probably the reasons for these differences. We aimed to sought clinical and pathological correlation that justifies the different clinical outcomes among COVID-19 autopsies cases.

**Methods:** Minimally invasive autopsy was performed on forty-seven confirmed COVID-19 patients from May-July, 2020. Electronic medical record of all patients was collected and a comprehensive histopathological evaluation was performed. Immunohistochemistry, immunofluorescence, special stain, western blotting and post-mortem real-time reverse transcriptase polymerase chain reaction on fresh lung tissue were performed.

**Results:** We show that 5/47 (10,6%) patients present a progressive decline in oxygenation index for acute respiratory distress syndrome (PaO_2_/FiO_2_ ratio), low compliance levels, interstitial fibrosis, high α-SMA+ cells/protein expression, high collagens I/III deposition and NETs(P<0.05), named as fibrotic phenotype (N=5). Conversely, 10/47 (21,2%) patients demonstrated progressive increase in PaO_2_/FiO_2_ ratio, high pulmonary compliance levels, preserved elastic framework, increase thrombus formation and high platelets and D-dimer levels at admission (P<0.05), named as thrombotic phenotype. While 32/47 (68,1%) had a mixed phenotypes between both ones.

**Conclusions:** We believe that categorization of patients based on these two phenotypes can be used to develop prognostic tools and potential therapies since the PaO_2_/FiO_2_ ratio variation and D-dimer levels correlate with the underlying fibrotic or thrombotic pathologic process, respectively; which may indicate possible clinical outcome of the patient.

## INTRODUCTION

The Severe Acute Respiratory Syndrome Coronavirus-2 (SARS-CoV-2) was first reported in Wuhan in China in 2019^1^. Infected patients can experience the less severe typical flu-like symptoms with fever, dry cough, and dyspnea^2^. However, in more severe cases, patients require intensive care unit (ICU) admission and mechanical ventilatory support. Acute respiratory distress syndrome (ARDS) is a major complication for severely infected patients and can lead to multi-organ failure such as acute kidney injury, cardiac complications and thromboembolic events ^3 4^. Differences in these outcomes can be attributed to a differential host response to infection with variable viral load, age, gender, comorbidities, genetic and immune background. While different studies have stratified heterogenous COVID-19 patients based on ABO blood grouping^5^, cancer subtypes^6^, hyper-inflammatory ^7,8^, the characterization of histopathological phenotypes in COVID-19 patients could reveal the underlying pathophysiological process, but it remains largely unreported.

Studies describing the clinical progression of patients who died from COVID-19 correlated to their histopathological pattern are still lacking. We hypothesized that SARS-CoV-2 infection can lead to different mechanisms of lung injury/repair with different clinical and ventilatory manifestations, even though they all lead to patient death. Therefore, we aimed to assess whether there is a clinical and pathological basis that could explain the different clinical outcomes among COVID-19 patients with commonly associated long-term lung dysfunction. Here we present a unique histopathologic analysis of COVID-19 autopsy cases with clinical and pathological correlation and demonstrate a distinct bimodal phenotypic characterization.

## METHODS

### Study Design

Forty-seven consecutive COVID-19 patients with positive nasopharyngeal swab for SARS-CoV-2 by reverse transcription polymerase chain reaction (RT-PCR) were considered eligible for this study. Modified minimally invasive autopsy (MIA)^9^ was performed at University Hospital of Ribeirão Preto Medical School, University of São Paulo, Ribeirão Preto, SP, Brazil – HCFMRP/USP from May to July, 2020. Briefly, all MIAs were performed at bedside through post-mortem surgical lung biopsy within 1 hour of death by a 3 cm incision on the anterior side of the chest between the fourth and fifth ribs. A matching 14-gauge cutting needle (Magnum Needles, Bard) and a biopsy gun (Magnum, Bard) also were used. Two post-COVID-19 biopsies cases were included. This study was approved by the local Research Ethics Committee and written informed consent was waived.

### Data Collection

Electronic medical record of all patients (N=47) enrolled in this study was collected. Demographic, clinical data, symptoms, drugs, treatments prescribed and laboratory tests were recorded thoroughly. Laboratory tests were collected and identified as: 1) Admission tests: up to the first 48 hours of admission; and 2) Daily tests: on the fifth, third and first day before death.

In order to better understand patient’s outcome, mechanical ventilation parameters of each individual patient were recorded. All daily change of these variables (from the first to the last day of mechanical ventilation until death) were registered and the means recorded for the last day before death of each individual. In addition, PaO_2_/FiO_2_ linear regression was performed with all available values. Then, to determine which patients had changes of lung function, we calculated the larger and smaller angles from the lines of PaO_2_/FiO_2_ linear regression. Finally, chest X-ray (CXR) and high-resolution computed tomography (HRCT) images of all patients were reviewed and evaluated by two specialized pulmonary radiologists (MKS and DTW). Unfortunately, only 15 patients had HRCT scan, preventing further analyses.

### Histological Evaluation

Lung tissue samples were fixed in 10% buffered formalin, for at least 48 h. Paraffin-embedded sections of 3-μm thickness were stained with Haematoxylin and Eosin, Picrosirius red (Abcam, ab150681) for collagen fibers identification and Verhoeff (Abcam, ab150667) for elastic fibers identification, according to local protocol. Immunohistochemistry for anti-alpha smooth muscle actin (α-SMA) (Abcam, ab5694) and anti-SARS-CoV-2 polyclonal antibody, developed by our group for in situ detection of SARS-CoV-2, were performed in paraffin-embedded sections of 3-μm thickness, following our lab protocol^10^. Histological evaluation was performed by specialized pulmonary pathologists (MLB, VLC, ATF) blinded to clinical history. For all patients, histopathological features were assessed as present or absent. Cut-offs of area involvement were determined for fibrotic phenotype, OP and AFOP, while thrombus formation was considered present or absent.

### Histomorphometry

Histochemistry and immunohistochemistry stains were quantified by morphometry. The images were captured with a digital camera on microscope (Novel L3000 LED) and analyzed using Image Pro Plus 7 software. Quantification followed the morphometric standards established by American Thoracic Society and European Thoracic Society (ATS/ERS)^11^. Positive α-sma cells were evaluated in ten different, randomly selected high-power fields of the lungs. At 400x magnification, the number of positive cells in each field was calculated according to the number of points hitting positive cells as a proportion of the total grid area. The density of collagen and elastic fibers was measured in the lung parenchyma in ten randomly selected microscopic fields at a magnification of ×200. The threshold for collagen fibers was established for all slides after the contrast was enhanced to the point at which the fibers were easily identified as green or orange bands. The density of the collagen and elastic fibers was expressed as the ratio between the measured fibers divided by the total area studied ×100.

### Immunofluorescence Analysis

Neutrophils and neutrophil-derived extracellular traps (NETs) were identified and quantified by immunofluorescence with anti-myeloperoxidase (MPO) (Abcam, ab25989), anti-citrullinated histone H3 (H3Cit) (Abcam, ab5103) and DAPI (Life Technologies, D1306). The staining and quantification were performed as our lab protocol^12^.

### Western Blotting Analysis

Protein extraction were performed for all fresh lung samples using QIAzol Lysis Reagent (Qiagen), according to our standered protocol. Protein levels were measured with the Bio-Rad Protein Assay (cat. 5000006) and transferred into a nitrocellulose membrane (GE Healthcare Biosciences, cat. 10600002). The membranes were incubated with anti-α-SMA (Abcam, ab7817), anti-matrix metalloproteinase-2 (anti-MMP-2) (Abcam, ab97779) and anti-glyceraldehyde-3-phosphate dehydrogenase (anti-GAPDH) (Abcam, ab9485). The proteins were detected and quantified with ChemiDoc Imaging Systems (Bio-Rad) and the results were normalized by GAPDH expression.

### Real-Time Polymerase Chain Reaction for Viral RNA

Total RNA from fresh lung tissue of all patients was obtained using QIAzol Lysis Reagent (Qiagen). Reverse transcriptase was performed using SuperScript IV reverse transcriptase (Invitrogen, Carlsbad, USA) according to manufacturer’s instructions and our standard protocol. RT-PCR for detection of SARS-CoV-2 nucleocapsid (N2) and envelope (E) viral proteins was performed on Step One Plus Real-Time PCR System (Applied Biosystems,Grand Island, NY) with TaqMan probes (Applied Biosystems). Both primers were designed by ThermoFisher Scientific from N2 and E genes sequences developed by Centers for Disease Control and Prevention (CDC) of Atlanta (USA)^13^ and Charité Germany^14^, respectively. Samples were tested in triplicate and cycle threshold values less than 44 were considered positive.

### Statistical Analysis

Statistical analysis was performed with SPSS v.13.0 0 software (SPSS, Inc., Chicago, IL, 2004). The data was evaluated using T test. Furthermore, nonparametric correlation (Spearman) was also performed with all collected data. Linear regression was also performed for these patients with all available PaO_2_/FiO_2_ values. Data are expressed as mean ± standard deviation and the p value less than 0.05 was be considered statistically significant.

## RESULTS

### COVID-19 Autopsy Cases: Demographic, Clinical, and Imaging Data

47 COVID-19 cases were confirmed by RT-PCR from nasopharyngeal swab. Following diagnosis, we conducted minimally invasive autopsies (MIA) on all positive patients at our institution (HCFMRP/USP). Demographic and clinical data for the 47 COVID-19 patients are summarized in Table 1.

**Table 1 –.** COVID-19 Clinical Data.

| All patients (N=47) |  |
| --- | --- |
| **DEATH AS A STARTING POINT** |  |
| Death after onset of COVID-19 symptoms (days)* | 18.90 ± 11 |
| Hospitalization (days)* | 14.43 ± 10.13 |
| **RT-PCR** |  |
| N2 gene (copies/mg)* | 8.81x10<sup>6</sup> ± 26.3x10<sup>6</sup> |
| E gene (copies/mg)* | 2.83x10<sup>8</sup> ± 7.65x10<sup>8</sup> |
| **DEMOGRAPHICS** |  |
| Gender (M;F) | 24;23 |
| Age (yrs)* | 67.83 ± 15.14 |
| Height (cm)* | 1.68 ± 0.10 |
| Weight (kg)* | 87.46 ± 28.53 |
| Body mass index (Kg/m<sup>2</sup>)* | 31.19 ± 9.08 |
| **COMORBIDITIES** |  |
| Systemic arterial hypertension (%) | 26 (55.3) |
| Obesity (%) | 17 (36.2) |
| Smokers (%) | 14 (29.8) |
| Diabetes (%) | 10 (21.3) |
| Chronic cardiovascular disease (%) | 10 (21.3) |
| Chronic respiratory disease (%) | 10 (21.3) |
| Chronic renal disease (%) | 7 (14.9) |
| Alcoholism (%) | 4 (8.5) |
| Others (%) | 40 (85.1) |
| **FIRST SYMPTOMS AND SIGNS IDENTIFIED** |  |
| Cough (%) | 24 (51.1) |
| Fever (%) | 16 (34.0) |
| Dyspnea (%) | 17 (36.2) |
| Flu-like symptoms (%) | 5 (10.6) |
| Myalgia (%) | 10 (21.3) |
| **SYMPTOMS AND SIGNS BEFORE ADMISSION** |  |
| Cough (%) | 16 (34.0) |
| Fever (%) | 9 (19.1) |
| Dyspnea (%) | 31 (66.0) |
| Flu-like symptoms (%) | 2 (4.3) |
| Myalgia (%) | 4 (8.5) |
| **SYMPTOMS AND SIGNS ON ADMISSION** |  |
| Cough (%) | 18 (38.3) |
| Fever (%) | 5 (10.6) |
| Dyspnea (%) | 37 (78.7) |
| Flu-like symptoms (%) | 0 (0.0) |
| Myalgia (%) | 0 (0.0) |
| **SYMPTOMS AND SIGNS DURING HOSPITALIZATION** |  |
| **Cough (%)** | 18 (38.3) |
| **Fever (%)** | 14 (29.8) |
| **Dyspnea (%)** | 19 (40.4) |
| **Flu-like symptoms (%)** | 0 (0.0) |
| **Myalgia (%)** | 0 (0.0) |
| **NEUROLOGICAL SYMPTOMS** |  |
| **Neurological symptoms (%)** | 7 (14.9) |
| **Headache (%)** | 4 (8.5) |
| **Dysgeusia (%)** | 3 (6.4) |
| **Anosmia (%)** | 5 (10.6) |
| **CLINICAL COMPLICATIONS** |  |
| **Septic shock (%)** | 29 (61.7) |
| **Acute kidney injury (%)** | 24 (51.1) |
| **Acute respiratory distress syndrome (%)** | 21 (44.7) |
| **Myocardial infarction (%)** | 12 (25.5) |
| **Congestive heart failure (%)** | 4 (8.5) |
| **MAIN MEDICATIONS** |  |
| **Anticoagulant (%)** | 47 (100.0) |
| **Steroids (%)** | 38 (80.9) |
| **Vasopressor (%)** | 37 (78.7) |
| **Chloroquine/Hydroxychloroquine (%)** | 2 (4.3) |
| **LABORATORY TESTS: ADMISSION (Highest value)** |  |
| **Lactate dehydrogenase (U/L)\*** | 783.12 ± 686.64 |
| **C-reactive protein (mg/L)\*** | 13.87 ± 8.72 |
| **D-dimer (mg/L)\*** | 4.22 ± 3.85 |
| **TREATMENTS** |  |
| **Prone positioning (%)** | 28 (59.6) |
| **Neuromuscular blockade (%)** | 29 (61.7) |
| **MECHANICAL VENTILATION PARAMETERS** |  |
| **Time of death after mechanical ventilation (days)\*** | 14.53 ± 10.70 |
| **Mechanical Ventilation (%)** | 36 (76.6) |
| **PaO<sub>2</sub>/FiO<sub>2</sub> ratio\* †** | 172.22 ± 90.64 |
| **Drive pressure cmH<sub>2</sub>O \* †** | 13.70 ± 4.42 |
| **Plateau pressure cmH<sub>2</sub>O \* †** | 22.85 ± 4.18 |
| **PICO cmH<sub>2</sub>O\* †** | 29.50 ± 8.18 |
| **PEEP cmH<sub>2</sub>O \* †** | 9.27 ± 2.21 |
| **Compliance ml/cmH<sub>2</sub>O\* †** | 31.53 ± 15.53 |

**Note:** \* mean ± standard deviation; (%) percentage; † Last measure before death.

The mean time from the onset of symptoms to death was 18.9 days with a mean hospital length of stay of 14.4 days. Patients were equally distributed by gender and the mean age was 67.8. Preexisting medical comorbidities were present in all patients, with systemic arterial hypertension as the most common (55%) followed by obesity (36%). The most frequent initial symptom was cough (55%). Dyspnea was present before the admission (66%), at the time of admission (79%), and during the hospital stay (40%). Clinical complications post hospital admission includes septic shock (62%), acute renal failure (51%) and acute respiratory distress syndrome (ARDS) (45%).

Retrospective review of chest X-ray (CXR) and high-resolution computed tomography (HRCT) of the chest was performed for all patients. On hospital admission, ground-glass opacities (GGO) were found in all 47 cases, and interstitial opacities were noted in 23 cases (48.9%). Consolidations were present in 13 of 15 cases (87%). Additional findings include traction bronchiectasis (N=6; 40%) and architectural distortion (N=8; 53.3%) (Supplementary Table 1).

Retrospective review of in-hospital treatments includes the use of steroids (81%) and prophylactic anticoagulation therapy (100%). Initial laboratory tests, collected in the first 48 hours post admission, demonstrated increased values of lactate dehydrogenase, C-reactive protein and D-dimer (Table 1). Additional laboratory tests are described in Supplementary Table 2. Mechanically ventilated patients (N=36, 76.6%), received neuromuscular blockade and prone positioning treatment (Table 1). PaO_2_/FiO_2_ ratio statistically correlated with D-dimer levels (r=0.37; P<0.05) (Supplementary Fig. 1A).

### Covid-19 Bimodal Clinical and Pathological Phenotypes

The PaO_2_/FiO_2_ linear regression, based on daily measure from the last value recorded for PaO_2_/FiO_2_ (before death) to the onset of mechanical ventilation, showed a bimodal clinical-pathological phenotype (Table 2):

**Table 2 -.** Bimodal Clinical and Pathological Phenotypes: Mechanical Ventilation Data.

| **MECHANICAL VENTILATION PARAMETERS** | **Thrombotic Phenotype** | **Fibrotic Phenotype** | **P** |
| --- | --- | --- | --- |
| **Time of death after mechanical ventilation (days)\*** | 11.4 ± 5.97 | 10.0 ± 4.53 | 0.65 |
| **Mechanical Ventilation (%)** | 10 (100.0) | 5 (100.0) | 0.99 |
| **Δ PaO<sub>2</sub>/FiO<sub>2</sub> \*** | 141.0 ± 201.0 | -169.50 ± 99.0 | 0.01 |
| **PaO<sub>2</sub>/FiO<sub>2</sub> ratio\* †** | 198.39 ± 90.25 | 101.16 ± 39.31 | 0.01 |
| **Drive pressure cmH<sub>2</sub>O\* †** | 12.22 ± 3.53 | 18.08 ± 4.70 | 0.06 |
| **Plateau pressure cmH<sub>2</sub>O\* †** | 21.38 ± 4.0 | 25.75 ± 3.30 | 0.12 |
| **PICO cmH<sub>2</sub>O\* †** | 25.3 ± 10.0 | 31.0 ± 5.57 | 0.57 |
| **PEEP cmH<sub>2</sub>O\* †** | 9.78 ± 1.56 | 8.50 ± 1.91 | 0.31 |
| **Compliance ml/cmH<sub>2</sub>O\* †** | 41.42 ± 16.32 | 19.80 ± 6.17 | 0.02 |

**Note:** \* mean ± standard deviation; (%) percentage; Δ Variation between the last (before death) and first (onset of mechanical ventilation) value recorded for PaO<sub>2</sub>/FiO<sub>2</sub>; † Last measure before death

1. Fibrotic Phenotype (N=5) – characterized by progressive decline in PaO_2_/FiO_2_ ratio and low compliance levels during hospitalization (Figure 1A); and alveolar septal thickening with myxoid fibrosis typical to an organizing-phase of diffuse alveolar damage (DAD) (Figure 1C; Table 3). Compliance correlated negatively with drive pressure for fibrotic phenotype (r=-0.67; P<0.05) (Supplementary Fig. 1C).
2. Thrombotic Phenotype (N=10) – characterized by a progressive increase in PaO_2_/FiO_2_ ratio and high pulmonary compliance levels during hospitalization (Figure 1B); and resolution of the acute/subacute lung injury to near to normal or slight distortion of the underlying lung parenchyma architecture (Figure 1D; Table 3), and with high frequency of thrombosis (Figure 1D).

**Figure 1 –.**
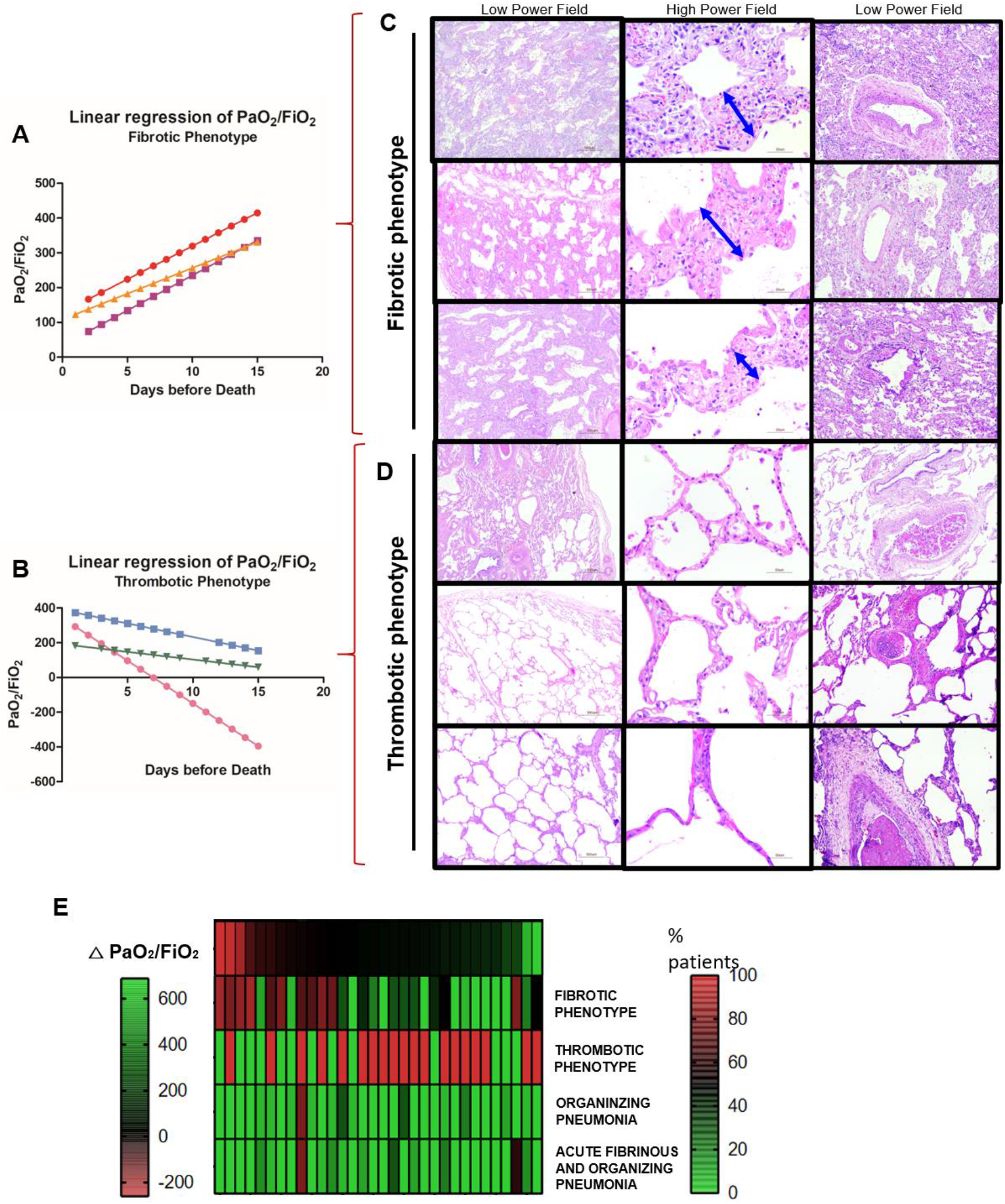
Bimodal Clinical and Histopathological Phenotypes. PaO_2_/FiO_2_ linear regression is based on daily measurement from the last value recorded (before death) to the onset of mechanical ventilation. It correlates with distinct histopathological features showing a bimodal phenotype: 1) Fibrotic Phenotype (N=5) showing progressive decline in PaO_2_/FiO_2_ ratio **(A)**, and significant alveolar septal thickening by fibrosis **(C** – double blue arrows, **E)** without thrombus formation **(C)**; and 2) Thrombotic Phenotype (N=10) showing progressive increase in PaO_2_/FiO_2_ ratio **(B)**, and resolution of acute/subacute lung injury to normal or near to normal parenchyma architecture and thrombus formation on vessels **(D, E)**. A population showing a mixed phenotype of lung injury (N=31) with morphologic features from both fibrotic and thrombotic phenotypes, including but not limited to organizing pneumonia, acute and fibrinous organizing pneumonia, and variable amounts of interstitial scarring, was present. This population seems to represent an intermediate non-bimodal phenotype, showing involvement by different stages of acute, organizing, and fibrotic DAD, that appears to fall between two opposite ends of lung injury manifesting as fibrotic and thrombotic respectively. Interestingly, a moderate to low frequency of thrombosis was noticed in these patients **(E)**.

**Table 3 -.** Bimodal Clinical and Pathological Phenotypes: Histopathological Findings in Minimally Invasive Autopsy.

| **HISTOLOGICAL FEATURES** | **Thrombotic Phenotype** | **Fibrotic Phenotype** | **P** |
| --- | --- | --- | --- |
| **Fibrotic septal thickening (>20% area)** | 4 (40.0) | 5 (100.0) | 0.01 |
| **Organizing pneumonia (>5% area)** | 2 (20.0) | 0 (0.0) | 0.52 |
| **Acute fibrinous and organizing pneumonia (>5% area)** | 1 (10.0) | 0 (0.0) | 0.99 |
| **Thrombus formation (%)** | 8 (80.0) | 2 (4.0) | 0.25 |

**Note:** (%) percentage.

Clinical and radiological data from both phenotypes are described in Supplementary Tables 4 to 6. D-dimer and platelets count was significantly higher in patients with thrombotic phenotype compared to fibrotic phenotype (Supplementary Fig. 2A, B and Supplementary Table 5; P <0.05). Findings from CRX image study showed similar distribution between bimodal phenotypes (Supplementary Table 6).

### COVID-19 Mixed Phenotype

A detailed histomorphological review of 47 MIA was performed to better understand COVID-19 induced lung injury (Supplementary Table 3). Variable amounts of alveolar septal thickening by fibrosis (interstitial fibrosis) were identified as the most common feature affecting the majority of the patients (N=41; 87.2%) (Supplementary Table 3). These findings significantly correlated with PaO_2_/FiO_2_ ratios (r=-0.42; P<0.05) (Supplementary Fig. 1B). In our cohort of 47 patients, we observe a unique bimodal phenotype in 15 patients (32%). However, 32 patients (68%) revealed a mixed phenotype of lung injury showing morphologic features from both fibrotic and thrombotic phenotypes. This population seems to represent an intermediate non-bimodal phenotype that appears to fall between two opposite ends manifesting as fibrotic and thrombotic respectively (Figure 1E). They may represent an intermediate phenotype showing different stages of involvement acute/subacute lung injury and fibrotic DAD morphologic features including organizing pneumonia (OP), acute fibrinous and organizing pneumonia (AFOP) with intra-alveolar fibrin “balls”, and variable amounts of interstitial scarring. Interestingly, a moderate to low frequency of thrombosis was noticed in these patients (Figure 1E).

### Pathological features of Covid-19 Fibrotic Phenotype

The fibrotic phenotype demonstrated in our cohort appears to be triggered by an imbalance between degradation and production of extracellular matrix (ECM) by myofibroblastic activation. Severe decrease in ECM elastic fibers was found in lungs from patients presenting the fibrotic phenotype, contrasting with the preserved elastic framework seen in lungs from patients with the thrombotic phenotype, as highlighted by Verhöeff staining (P<0.05) (Figure 2A, B). This is further supported by the presence of a direct association between the area fraction of elastic fibers in ECM and the compliance levels during mechanical ventilation (r=0.57; P<0.05) (Supplementary Fig. 1D). The fibrotic phenotype patients showed prominent accumulation of α-SMA in ECM, a marker of myofibroblast activity and neovasculogenesis, compared to thrombotic phenotype patients (P <0.05) (Figure 2A, C). There was also an inverse association between the area fraction of elastic fibers and myofibroblasts in ECM (r=-0.57; P<0.05) (Supplementary Fig. 1E). α-SMA and anti-MMP-2 protein expression by Western blot were increased in the fibrotic phenotype (P<0.05) (Figure 2D), reinforcing the fibrotic nature of this process.

**Figure 2 –.**
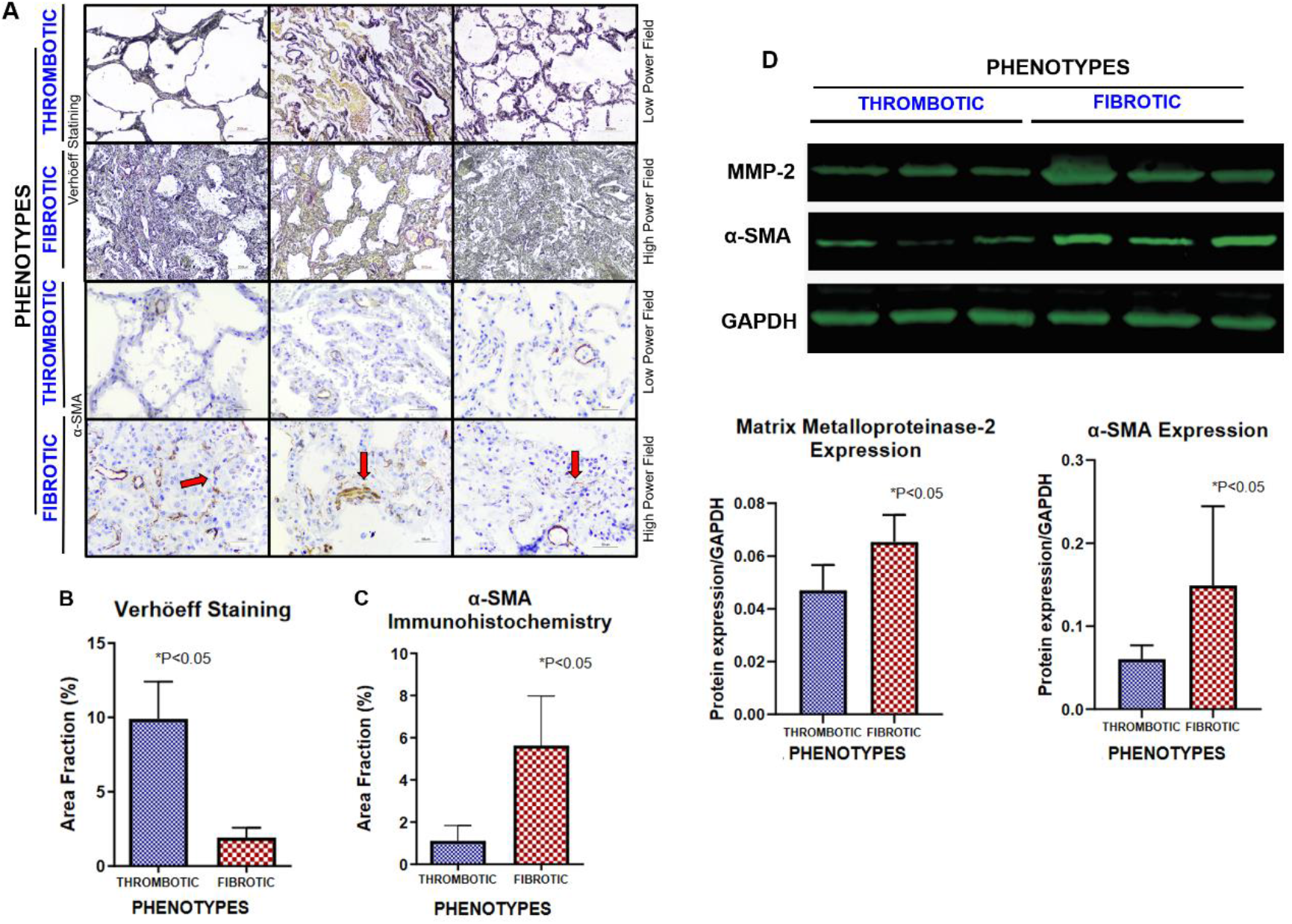
Bimodal Clinical and Pathological Phenotypes: Elastic Fibers Framework and Myofibroblastic Activation. Fibrotic phenotype demonstrates higher levels of α-SMA expression (red arrows), indicating extracellular matrix deposition by myofibroblasts and neovasculogenesis, resulting in lung tissue scarring and stiffening (P <0.05) **(A** – red arrow, **C)**. Consequently, lung elastic fibers related to tissue elastic capacity (% area fraction of elastolysis), was significantly reduced in fibrotic phenotype when compared to thrombotic phenotype (P <0.05) **(A, B)**. The cicatricial nature of the process is further supported by Western blot, revealing higher protein expression levels of α-SMA and MMP-2 in fibrotic phenotype **(D)**. GAPDH was used as loading control for gene expression.

Picrosirius Red staining demonstrated a significant increase in ECM deposition by green-birefringence thin fibers (most by types III collagen) and red/orange-birefringence thick fibers (most by type III collagen) in lungs from patients with fibrotic phenotype (P<0.05) (Figure. 3B, C). These findings significantly correlate with PaO_2_/FiO_2_ ratios (r=-0.64; P<0.05) (Supplementary Fig. 1F). Additionally, neutrophils with significantly higher length of NETs were also observed in these patients (P<0.05) (Supplementary Fig. 3A, C).

**Figure 3 –.**
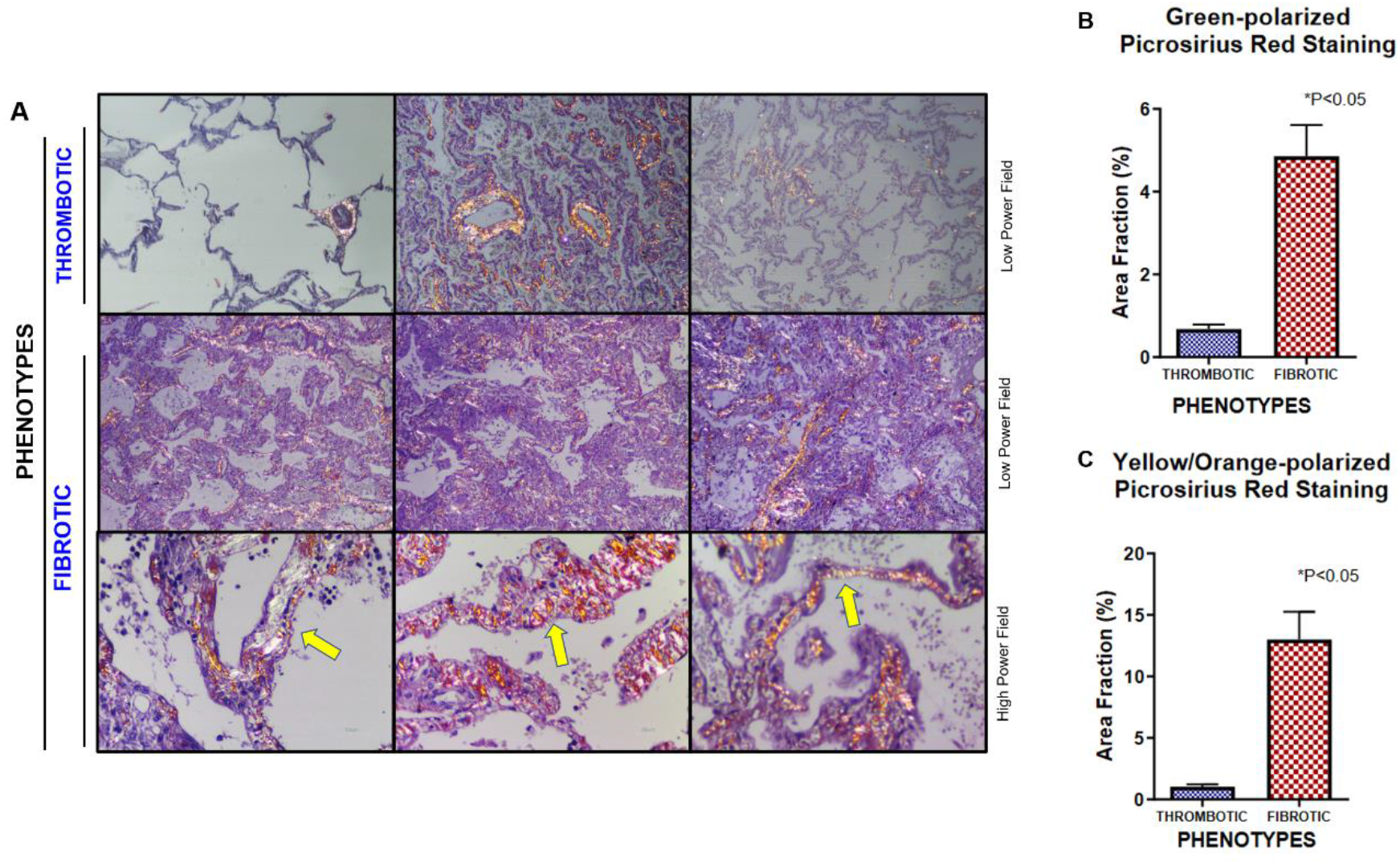
Bimodal Clinical and Pathological Phenotypes: Collagen Depositions. Extensive collagen production and interstitial deposition was noted, characterized by the presence of alveolar septal expansion by dense fibrosis (yellow arrows), which was more prominent in the fibrotic phenotype, confirmed by picrosirius red staining **(A** – yellow arrow). Area fraction percentage of collagen fibers labeled by green and yellow orange-polarized picrosirius red stain was significantly higher in fibrotic phenotype compared with thrombotic one, as demonstrated by P <0.05 **(B, C)**.

### Fibrotic Phenotype in Post-COVID-19 Survivors Follow Up

Post COVID-19 infection follow up with transbronchial biopsies were performed at our institution (HCFMRP-USP) in two patients, both in their 60’s and male, ranging from three- to five-months’ post infection. Histopathologic analysis revealed minimal chronic inflammation with remodeling of airway wall by submucosal scarring and alveolar septal thickening by fibrosis (Figure 4). While this may represent scarring secondary to resolving infection, the possibility of future post-infection chronic fibrosing disease needs to be excluded. Secondly, more extensive studies are needed to determine if post COVID-19 complications arise in patients who survive this disease.

**Figure 4 –.**
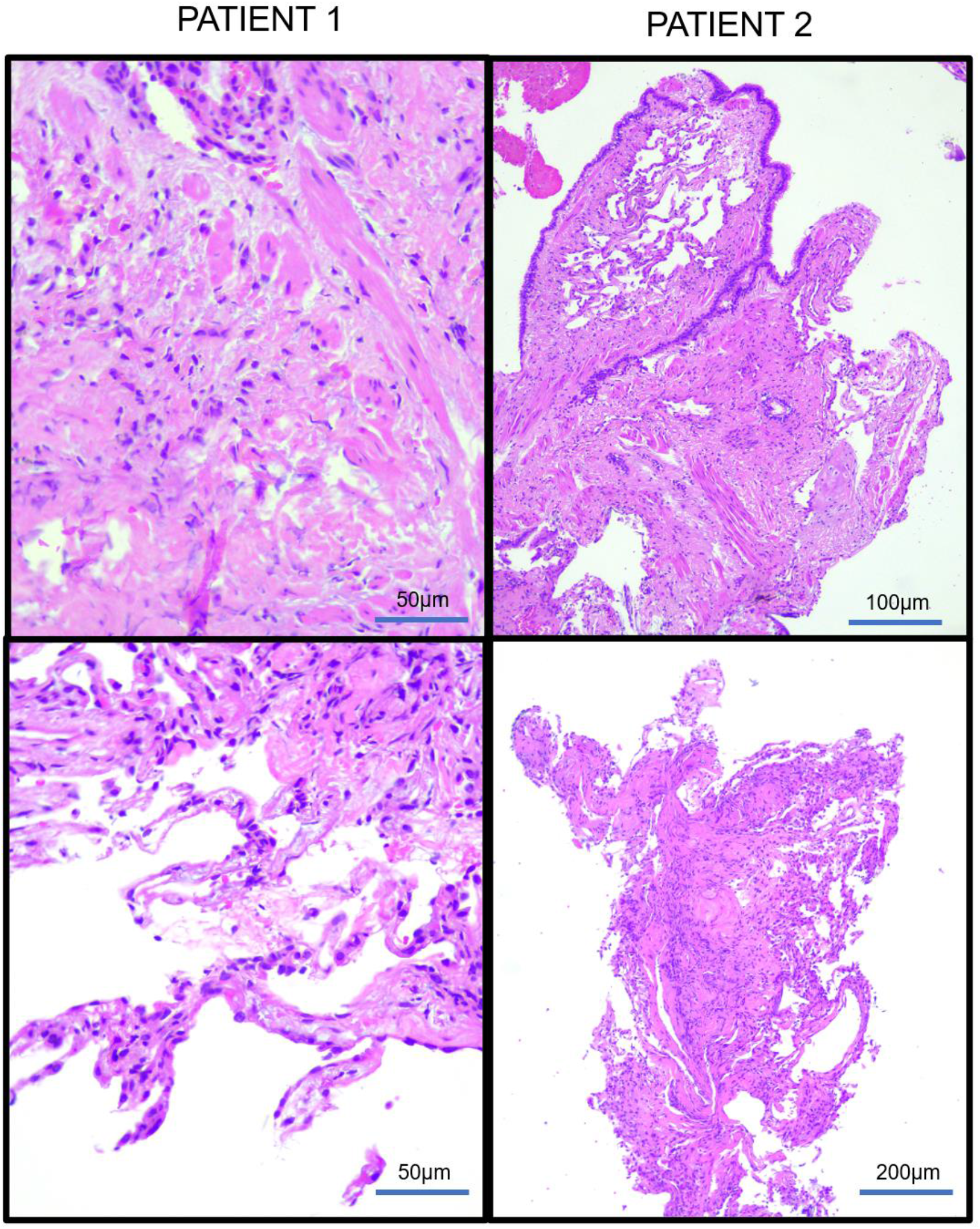
Post COVID 19 Infection Transbronchial Biopsies Follow Up. Two patients, both in their 60’s and male, ranging from three- to five-months’ post infection: There is minimal chronic inflammation and airway wall and lung parenchyma remodeling, characterized by submucosal scarring and alveolar septal thickening by fibrosis associated with alveolar pneumocyte prominence, respectively. While this may represent scarring secondary to resolving infection, the possibility of an evolving post infection chronic fibrosing disease needs to be excluded and more extensive studies are needed to investigate post COVID-19 complications in patients who survive the disease.

## DISCUSSION

We discovered two distinctive histopatholgical bimodal phentoypes and correlated histopathologic findings to corresponding clinical outcomes. Patients presenting with confirmed lung fibrosis predominantly displayed either thrombotic or fibrotic characterization, which correlated to differing clinical outcomes. This is in contrast to the initial pathophysiological events that occur in the acute phase of ARDS following post-viral insult to the lungs where the clinical outcomes diverge as infection progresses.

Upon viral insult to the airways and bronchial epithelial cells, a severe lymphocytic bronchiolitis occurs, promoting epithelial injury and cell death, eventually resulting in structural disarray by airway-centered remodeling (Figure 5). The aggressive airway wall infection then spreads to the lung parenchyma, inducing cellular pneumonitis, and damages the alveolar-capillary barrier (Figure 5, yellow arrow). This promotes the denudation of the alveolar septa, characterized as DAD, bronchiolocentric alveolar hemorrhage or fibroblastic plug, called OP, associated or not with fibrin balls, known as AFOP (Figure 5). Subsequently, two different outcomes of repair appear to occur depending on resolution or progression of the lung injury, eventually manifesting as a thrombotic phenotype with gradual recovering of the acute/subacute lung injury associated with thrombosis; or a fibrotic phenotype with sustained myofibroblastic proliferation with interstitial scarring and parenchymal remodeling (Figure 1, 5).

**Figure 5 –.**
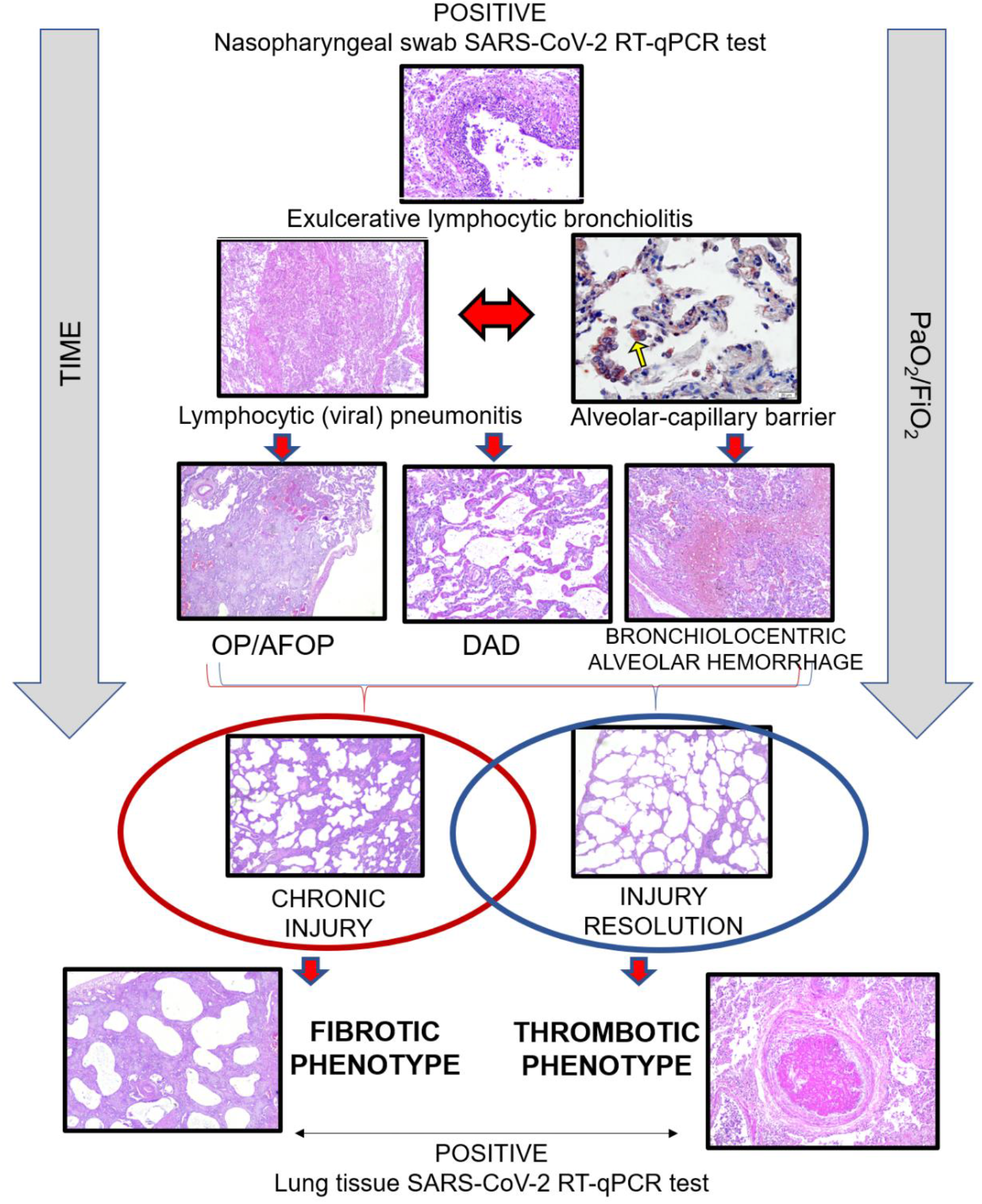
COVID-19 Lung Pathophysiology. Following bronchial wall viral infection there is severe lymphocytic bronchiolitis, promoting striking epithelial injury, cell loss, and mucosal ulceration, eventually resulting in airway remodeling. The aggressive airway wall infection then spreads to lung parenchyma, inducing cellular pneumonitis and damage of alveolar-capillary barrier (immunohistochemical stain decorates SARS-CoV-2 infected cells, yellow arrow). This promotes bronchiolocentric alveolar hemorrhage, acute DAD, and fibrin balls associated with fibroblastic plugs of organizing pneumonia, known as acute fibrinous and organizing pneumonia. Subsequently, two different ultimate ends of repair appear to occur depending on resolution or progression of the lung injury, eventually manifesting as a thrombotic phenotype, with gradual recovering of the acute/subacute lung injury associated with prominent thrombosis; or a fibrotic phenotype with sustained myofibroblastic proliferation, which promotes interstitial scarring and parenchymal remodeling.

The fibrotic phenotype seen in some patients is the end are the result of the activation and termination of many different pathways, one of which is collagen fibers deposition (type I and III) to cause alveolar septal thickening (Figure 3). Additionally, the fibrotic phenotype revealed higher active myofibroblasts α-SMA area fraction (Figure 2A, C) and higher α-SMA and MMP-2 expression (Figure 2D) (P <0.05) than the thrombotic phenotype. Similar studies also describes COVID-19 organizing DAD with α-SMA+ myofibroblasts^15^ and fibrous parenchyma remodeling^16,17^. Alveolar septal thickening by fibrosis impairs adequate lung function and gas exchange, which is reflected in our study by significant correlation between collagen fibers deposition and PaO_2_/FiO_2_ ratio in the fibrotic phenotype (r=-0.64; P <0.05) (Supplementary Fig. 1F). Furthermore, lung elastic capacity, represented by elastic fibers area fraction (Verhoeff stain quatification) was significantly reduced (P<0.05) (Figure 2A, B). Moreover, fibrotic phenotype patients presented NETs in a significantly higher extent throughout lung parenchyma compared to thrombotic phenotype subjects (P<0.05) (Supplementary Fig. 3). As described by our group previously^12^, the formation of neutrophil extracelular traps (NETs), networks of extracellular fibers, primarily composed of DNA from neutrophils, binded with pathogens ^18^, is an important response to SARS-CoV-2 during the acute and a potential contributory factor to later fibrotic phase by amplify the chronic reparative phase. This result suggests a more prominent host inflammatory response in the fibrotic phenotype subjects with consequently greater damage to lung parenchyma, trigging a more aggressive reparative fibroblastic process, as observed in these patients. In accordance with the histological analysis, the clinical data also suggested a poor clinical outcome. These patients presented progressive decline in PaO_2_/FiO_2_ ratio (Figure 1A) (mean 101.16) and low compliance levels during hospitalization (mean 19.8 ml/cmH_2_O) (Table 2). In accordance to other studies, low ventilatory parameters were also observed^19,20^ and patients presented a median value of 129 PaO_2_/FiO_2_ ratio and 27 ml/cmH_2_O compliance, as described by Cummings M et al^21^,

Conversely, subjects from the thrombotic phenotype showed an unexpected progressive increase in PaO_2_/FiO_2_ ratio (Figure 1B) (mean 141) and high pulmonary compliance levels (mean 41.4 ml/cmH_2_O) (Table 2). Similarly, a nearly normal lung compliance has also surprised Gattinoni et al^22^ as their patients presented 50.2 ml/cmH_2_O, despite the severe hypoxaemia. However, better ventilatory parameters in our thrombotic phenotype subjects could be justified by the gradual recovering of the acute/subacute lung injury (Figure 1D) with increased area fraction of elastic fibers (Figure 2A, B). Nevertheless, sudden death occurred, probably due to higher frequency of pulmonary thromboembolism (Figure 1D, Table 3), as also described in other studies^23,24^. As expected, D-dimer levels and platelets count were significantly higher in these patients than in fibrotic phenotype subjects (Supplementary Fig. 2A, B and Supplementary Table 5). High D-dimer levels in COVID-19 patients has been recently related with thrombosis as an important prognostic biomarker of severity ^25,26^. In addition, platelets have been reported with a hyperactivated phenotype in COVID-19 patients, possibly related to hypercoagulation^8^. While it’s well known that thrombus formation is a feature of organizing DAD, it’s unclear why some COVID-19 patients develop a more frequently form thrombi.

In light of this, we observed two distinct bimodal histopathological phenotypes with their correlative clinical outcomes among the 15 COVID-19 (32%) evidenced with fibrosis. This may justify the differing clinical outcomes observed in out cohort, despite the same etiology and the convergence to their endpoints- mortality. Likewise, Gattinoni et al^3^ have also reported different behaviors throughout COVID-19 patients’ outcome and classified them into two phenotypes: Type L: low elastance; and Type H: high elastance. Thus, COVID-19 patients with different clinical presentations are frequently described. However, our work provides insight into the importance of histopathological screening among COVID19 autopsies correlated to their differing clinical outcomes, with a particular focus in the lung repair resolving phase of injury.

Besides those two crucial phenotypes, it is important to mention that 32 patients (68%) had a mixed pattern of lung injury appearing as a non-bimodal phenotype (Figure 1E). The pathophysiology of these patients probably represents different stages of viral lung injury, with both features of acute/subacute and fibrotic/organizing lung injury with variable degrees of interstitial scarring, OP and AFOP. Despite the importance of this histopathological finding, the correlation with clinical data did not highlight any relevance. On the other hand, the clinical and pathological correlation of our 15 patients evidenced two distinctive bimodal opposite COVID-19 phenotypes, manifesting as fibrotic predominant and thrombotic predominant, being a unique and important finding.

Therefore, the reported sudden deaths of COVID-19 patients in clinical improvement and the description of post-COVID-19 tomographic fibrotic changes may be justified by our findings. In fact, two COVID-19 patients’ follow up with transbronchial biopsies, performed at 3 to 5 months post infection, demonstrated lung parenchyma remodeling with alveolar septal thickening by dense fibrosis (Figure 4). Pathophysiological differences in the virus versus immunity interaction may cause different clinical outcomes in COVID-19 patients, as demonstrated in our study after 47 COVID-19 autopsies. Our described bimodal clinical and pathological phenotypes are critically important in the management of infected patients, given that a progressive drop in PaO_2_/FiO_2_ ratio or high platelets/D-Dimer levels are the clinical findings that may raise the clinical suspicion for one of the two outcome phenotypes at the beginning of the hospitalization, allowing a possible better therapeutic management.

## Data Availability

The data that support the findings of this study are available from the corresponding author upon reasonable request. Source data are provided with this paper.

## ACKNOWLEDGMENTS

The authors would like to thank the institutional support of the University Hospital of Ribeirão Preto Medical School, University of São Paulo, Ribeirão Preto, SP, Brazil – HCFMRP/USP. This work was supported by São Paulo Research Foundation (FAPESP) [2019/01517-3, 2019/19591-5 and 2020/13370-4].

## STUDY APPROVAL

The procedures followed in the study were approved by the National Ethics Committee – Brazil (CAAE: 32475220.5.0000.5440). The written informed consent was waived.

