## Supplementary Material for "COVID-19 Bimodal Clinical and Pathological Phenotypes"

*Sabrina S Batah[1]; Maíra N Benatti[2]; Li Siyuan[3]; Wagner M Telini[1]; Jamile Barbosa[1]; Marcelo B Menezes[2]; Tales R Nadai[4]; Keyla S G Sá[5]; Chirag M. Vaswani[6,7]; Sahil Gupta[7,8,9] Dario S Zamboni[5]; Danilo T Wada[10]; Rodrigo T Calado[10]; Renê D R Oliveira[11]; Paulo Louzada-Junior[11]; Maria Auxiliadora-Martins[12]; Flávio P Veras[13]; Larissa D Cunha[5]; Thiago M Cunha[13]; Rodrigo Luppino-Assad[14;] Marcelo L Balancin[15]; Sirlei S Morais[1]; Ronaldo B Martins[5]; Eurico Arruda[5]; Fernando Chahud[1]; Marcel Koenigkam-Santos[10]; Andrea A Cetlin[2]; Fernando Q Cunha[13]; Claudia dos Santos[7,16]; Vera L Capelozzi[15]; Junya Fukuoka[17;] Rosane Duarte-Achcar[18]; Alexandre T Fabro[1*]*

[1]Department of Pathology and Legal Medicine, Ribeirão Preto Medical School, University of São Paulo, Brazil; [2]Pulmonary Division, Department of Internal Medicine, Ribeirão Preto Medical School, University of São Paulo, Brazil; [3]Department of Surgery, Ribeirão Preto Medical School, University of São Paulo, Brazil; [4]Hospital Estadual de Bauru, Brazil; [5]Department of Cell and Molecular Biology and Pathogenic Bioagents, Ribeirão Preto Medical School, University of São Paulo, Brazil; [6]Department of Physiology, Temerty Faculty of Medicine, University of Toronto, Toronto, ON, CA; [7]Keenan Research Centre for Biomedical Science, St. Michael's Hospital, Toronto, ON, CA; [8]Institute of Medical Science, Temerty Faculty of Medicine, University of Toronto, 1 King's College Circle, Toronto, ON, CA; [9]Department of Critical Care Medicine, St. Michael's Hospital, Toronto, ON, CA; [10]Department of Medical Images, Hematology and Oncology, Ribeirão Preto Medical School, University of São Paulo, Brazil; [11]Division of Clinical Immunology, Emergency, Infectious Diseases and Intensive Care Unit, Ribeirão Preto Medical School, University of São Paulo, Brazil; [12]Division of Intensive Care Medicine, Department of Surgery and Anatomy, Ribeirão Preto Medical School, University of São Paulo; Brazil; [13]Department of Pharmacology, Ribeirão Preto Medical School, University of São Paulo, Brazil; [14]Department of Internal Medicine, Ribeirão Preto Medical School, University of São Paulo, Brazil; [15]Department of Pathology, Faculty of Medicine, University of São Paulo, Brazil; [16]Interdepartmental Division of Critical Care Medicine, University of Toronto, Toronto, ON, Canada; [17]Department of Pathology, Nagasaki University Graduate School of Biomedical Sciences, Nagasaki, Japan; [18]National Jewish Health, Department of Medicine, Pathology Division, Denver, Colorado, USA.

**SUPPLEMENTARY TABLES**

**Supplementary Table 1 - COVID 19 Radiological Features.**

| **All patients (N=47)** | |
|---|---|
| CHEST XR | |
| **Ground-glass opacity (%)** | 47 (100.0) |
| **Edema (%)** | 8 (17.0) |
| **Pulmonary consolidation (%)** | 9 (19.1) |
| **Pulmonary congestion (%)** | 3 (6.4) |
| **Pleural effusion (%)** | 7 (14.9) |
| **Interstitial opacities (%)** | 23 (48.9) |
| **Cardiomegaly (%)** | 31 (65.9) |

| **15 patients** | |
|---|---|
| CHEST CT | |
| **Crazy-paving (%)** | 5 (33.3) |
| **Consolidations (%)** | 13 (86.7) |
| **Architectural distortion (%)** | 8 (53.3) |
| **Traction bronchiectasis (%)** | 6 (40.0) |
| **Honeycombing (%)** | 1 (6.7) |
| **Pleural effusion (%)** | 9 (60.0) |

**Note:** (%) percentage.

**Supplementary Table 2 - COVID-19 Laboratory Tests.**

| All patients (N=47) | |
|---|---|
| **DAILY TESTS (Highest value)** | |
| **White blood cells count ($10^3$/µL)*** | 21.77 ± 11.35 |
| **Hemoglobin (g/dL)*** | 11.06 ± 1.93 |
| **Sodium (mmol/L)*** | 140.8 ± 7.46 |
| **Potassium (mmol/L)*** | 5.12 ± 1.27 |
| **Creatinine (mg/dL)*** | 2.45 ± 1.51 |
| **Lymphocytes ($10^3$/µl)*** | 1.49 ± 1.14 |
| **Platelets ($10^3$/µl)*** | 287.36 ± 143.93 |
| **Neutrophils ($10^3$/µl)*** | 18.40 ± 9.31 |
| **LABORATORY TESTS (1, 3, and 5 days before death)** | |
| **One day before death** | |
| **White blood cells count ($10^3$/µL)*** | 21.85 ± 12.29 |
| **Hemoglobin (g/dL)*** | 9.62 ± 2.07 |
| **Sodium (mmol/L)*** | 137.88 ± 7.79 |
| **Potassium (mmol/L)*** | 4.94 ± 1.08 |
| **Creatinine (mg/dL)*** | 2.41 ± 1.45 |
| **Lymphocytes ($10^3$/µl)*** | 1.37 ± 1.18 |
| **Platelets ($10^3$/µl)*** | 264.05 ± 152.44 |
| **Neutrophils ($10^3$/µl)*** | 18.60 ± 9.53 |
| **Three days before death** | |
| **White blood cells count ($10^3$/µL)*** | 17.14 ± 8.82 |
| **Hemoglobin (g/dL)*** | 10.40 ± 2.29 |
| **Sodium (mmol/L)*** | 139.43 ± 6.99 |
| **Potassium (mmol/L)*** | 4.65 ± 0.97 |
| **Creatinine (mg/dL)*** | 2.13 ± 1.56 |
| **Lymphocytes ($10^3$/µl)*** | 1.12 ± 0.87 |
| **Platelets ($10^3$/µl)*** | 253.36 ± 113.81 |
| **Neutrophils ($10^3$/µl)*** | 15.14 ± 7.61 |
| **Five days before death** | |
| **White blood cells count ($10^3$/µL)*** | 15.78 ± 7.46 |
| **Hemoglobin (g/dL)*** | 10.61 ± 1.85 |
| **Sodium (mmol/L)*** | 139.57 ± 6.86 |
| **Potassium (mmol/L)*** | 1.66 ± 1.27 |
| **Creatinine (mg/dL)*** | 2.12 ± 1.31 |
| **Lymphocytes ($10^3$/µl)*** | 1.04 ± 0.86 |
| **Platelets ($10^3$/µl)*** | 266.15 ± 130.50 |
| **Neutrophils ($10^3$/µl)*** | 14.33 ± 6.63 |

**Note:** * mean ± standard deviation.

**Supplementary Table 3 - COVID19 Minimally Invasive Autopsy Findings.**

| **All patients (N=47)** | |
|---|---|
| HISTOLOGIC FEATURES | |
| **Interstitial fibrosis (%)** | 41 (87.2) |
| **Organizing pneumonia (%)** | 21 (44.7) |
| **Acute fibrinous organizing pneumonia (%)** | 33 (70.2) |
| **Thrombus formation (%)** | 25 (53.2) |
| **Viral cytopathic effect (%)** | 24 (51.1) |
| **Squamous metaplasia (%)** | 13 (27.7) |
| **Alveolar hemorrhage (%)** | 29 (61.7) |
| **Hyaline membrane (%)** | 22 (46.8) |
| **Hyalinization (%)** | 18 (38.3) |
| **Intra-alveolar edema (%)** | 6 (12.8) |
| **Neutrophilic infiltrate (%)** | 8 (17.0) |
| **Occult lung cancer (%)** | 17 (36.2) |
| **Bronchiolization (%)** | 21 (44.7) |
| **Bronchiectasis** | 24 (51.1) |
| **Normal pulmonary parenchyma** | 41 (87.2) |
| **Pneumocyte desquamation** | 20 (42.6) |
| **Cellular bronchiolitis (%)** | 3 (6.4) |
| **Pleural thickening (%)** | 8 (17.0) |

**Note:** (%) percentage.

**Supplementary Table 4 – Bimodal COVID-19 Phenotypes: Demographical and Clinical Data.**

| DEMOGRAPHIC AND CLINICAL DATA | Thrombotic Phenotype | Fibrotic Phenotype | P |
|---|---|---|---|
| **DEATH AS A STARTING POINT** | | | |
| **Time of death from onset of COVID-19 symptoms (days)*** | 18.30 ± 4.30 | 18.00 ± 6.12 | 0.92 |
| **Hospitalization length (days)*** | 14.10 ± 6.40 | 13.60 ± 4.22 | 0.88 |
| **RT-PCR** | | | |
| **N2 gene (copies/mg)** | $1.21x10^5 \pm 2.64x10^5$ | $3.92x10^6 \pm 8.49x10^6$ | 0.09 |
| **E gene (copies/mg)** | $2.06x10^7 \pm 3.59x10^7$ | $8.19x10^7 \pm 1.61x10^8$ | 0.61 |
| **Demographics** | | | |
| **Gender (M;F)** | 5;5 | 1;4 | 0.58 |
| **Age (yrs)*** | 64.60 ± 12.25 | 70.60 ± 7.09 | 0.83 |
| **Height (cm)*** | 1.69 ± 0.07 | 1.70 ± 0.07 | 0.96 |
| **Weight (kg)*** | 93.88 ± 31.84 | 82.67 ± 11.24 | 0.81 |
| **Body mass index (Kg/m²)*** | 32.61 ± 9.16 | 28.87 ± 5.93 | 0.63 |
| **COMORBIDITIES** | | | |
| **Systemic arterial hypertension (%)** | 7 (70.0) | 3 (60.0) | 0.99 |
| **Obesity (%)** | 4 (40.0) | 2 (40.0) | 0.99 |
| **Smokers (%)** | 1 (10.0) | 3 (60.0) | 0.07 |
| **Diabetes (%)** | 6 (60.0) | 4 (80.0) | 0.60 |
| **Chronic cardiovascular disease (%)** | 3 (30.0) | 0 (0.0) | 0.50 |
| **Chronic respiratory disease (%)** | 3 (30.0) | 0 (0.0) | 0.50 |
| **Chronic renal disease (%)** | 2 (20.0) | 1 (20.0) | 0.99 |
| **Alcoholism (%)** | 1 (10.0) | 0 (0.0) | 0.99 |
| **Others (%)** | 9 (90.0) | 4 (80.0) | 0.99 |
| **FIRST SYMPTOMS AND SIGNS IDENTIFIED** | | | |
| **Cough (%)** | 4 (40.0) | 4 (80.0) | 0.28 |
| **Fever (%)** | 3 (30.0) | 2 (40.0) | 0.99 |
| **Dyspnea (%)** | 2 (20.0) | 1 (20.0) | 0.99 |
| **Flu-like symptoms (%)** | 2 (20.0) | 0 (0.0) | 0.52 |
| **Myalgia (%)** | 4 (40.0) | 2 (40.0) | 0.99 |
| **SYMPTOMS AND SIGNS BEFORE ADMISSION** | | | |
| **Cough (%)** | 1 (10.0) | 1 (20.0) | 0.99 |
| **Fever (%)** | 1 (10.0) | 1 (20.0) | 0.99 |
| **Dyspnea (%)** | 6 (60.0) | 2 (40.0) | 0.60 |
| **Flu-like symptoms (%)** | 1 (10.0) | 0 (0.0) | 0.99 |
| **Myalgia (%)** | 1 (10.0) | 0 (0.0) | 0.99 |
| **SYMPTOMS AND SIGNS ON ADMISSION** | | | |
| **Cough (%)** | 1 (10.0) | 1 (20.0) | 0.99 |
| **Fever (%)** | 1 (10.0) | 0 (0.0) | 0.99 |
| **Dyspnea (%)** | 10 (100.0) | 4 (80.0) | 0.33 |

| | | | |
|---|---|---|---|
| **Flu-like symptoms (%)** | 0 (0.0) | 0 (0.0) | 0.99 |
| **Myalgia (%)** | 0 (0.0) | 0 (0.0) | 0.99 |
| **SYMPTOMS AND SIGNS DURING HOSPITALIZATION** | | | |
| **Cough (%)** | 4 (40.0) | 2 (20.0) | 0.99 |
| **Fever (%)** | 6 (60.0) | 3 (60.0) | 0.99 |
| **Dyspnea (%)** | 3 (30.0) | 2 (20.0) | 0.99 |
| **Flu-like symptoms (%)** | 0 (0.0) | 0 (0.0) | 0.99 |
| **Myalgia (%)** | 0 (0.0) | 0 (0.0) | 0.99 |
| **NEUROLOGICAL SYMPTOMS** | | | |
| **Neurological symptoms (%)** | 2 (20.0) | 0 (0.0) | 0.52 |
| **Headache (%)** | 1 (10.0) | 0 (0.0) | 0.99 |
| **Dysgeusia (%)** | 0 (0.0) | 0 (0.0) | 0.99 |
| **Anosmia (%)** | 2 (20.0) | 0 (0.0) | 0.52 |
| **CLINICAL COMPLICATIONS** | | | |
| **Septic shock (%)** | 8 (80.0) | 4 (80.0) | 0.99 |
| **Acute kidney injury (%)** | 8 (80.0) | 4 (80.0) | 0.99 |
| **Acute respiratory distress syndrome (%)** | 4 (40.0) | 5 (100.0) | 0.88 |
| **Myocardial infarction (%)** | 4 (40.0) | 1 (20.0) | 0.60 |
| **Congestive heart failure (%)** | 1 (10.0) | 0 (0.0) | 0.99 |
| **MAIN MEDICATIONS** | | | |
| **Anticoagulant (%)** | 10 (100.0) | 5 (100.0) | 0.99 |
| **Steroids (%)** | 9 (90.0) | 5 (100.0) | 0.99 |
| **Vasopressor (%)** | 10 (100.0) | 4 (80.0) | 0.33 |
| **Chloroquine/Hydroxychloroquine (%)** | 1 (10.0) | 0 (0.0) | 0.99 |
| **TREATMENTS** | | | |
| **Prone positioning (%)** | 10 (100.0) | 3 (60.0) | 0.95 |
| **Neuromuscular blockade (%)** | 10 (100.0) | 4 (80.0) | 0.33 |

**Note:** * mean ± standard deviation; (%) percentage

**Supplementary Table 5 – Bimodal COVID-19 Phenotype: Clinical Data.**

| LABORATORY TESTS | Thrombotic Phenotype | Fibrotic Phenotype | P |
|---|---|---|---|
| **ADMISSION TESTS (Highest value)** | | | |
| **Lactate dehydrogenase (U/L)*** | 422.11 ± 169.09 | 645.30 ± 77.88 | 0.09 |
| **C-reactive protein (mg/L)*** | 16.76 ± 9.45 | 12.60 ± 9.52 | 0.67 |
| **D-dimer (mg/L)*** | 13.26 ± 6.27 | 2.99 ± 1.628 | 0.02 |
| **DAILY TESTS (Highest value)** | | | |
| **White blood cells count ($10^3$/µL)*** | 25.03 ± 10.36 | 27.18 ± 12.08 | 0.85 |
| **Hemoglobin (g/dL)*** | 11.99 ± 1.15 | 11.16 ± 1.04 | 0.24 |
| **Sodium (mmol/L)*** | 141.60 ± 6.23 | 137.50 ± 6.06 | 0.43 |
| **Potassium (mmol/L)*** | 5.19 ± 0.99 | 5.77 ± 0.94 | 0.30 |
| **Creatinine (mg/dL)*** | 3.57 ± 1.87 | 3.48 ± 1.10 | 0.99 |
| **Lymphocytes ($10^3$/µl)*** | 2.5 ± 1.41 | 1.40 ± 1.31 | 0.37 |
| **Platelets ($10^3$/µl)*** | 364.00 ± 129.07 | 225.60 ± 74.87 | 0.04 |
| **Neutrophils ($10^3$/µl)*** | 20.44 ± 8.58 | 24.28 ± 10.47 | 0.70 |
| **LABORATORY TESTS (1, 3, and 5 days before death)** | | | |
| **One day before death** | | | |
| **White blood cells count ($10^3$/µL)*** | 25.59 ± 11.29 | 27.23 ± 12.12 | 0.80 |
| **Hemoglobin (g/dL)*** | 10.31 ± 1.97 | 9.05 ± 1.59 | 0.36 |
| **Sodium (mmol/L)*** | 138.93 ± 7.40 | 131.25 ± 5.06 | 0.15 |
| **Potassium (mmol/L)*** | 4.80 ± 1.22 | 5.53 ± 0.90 | 0.28 |
| **Creatinine (mg/dL)*** | 3.47 ± 2.00 | 2.93 ± 1.12 | 0.68 |
| **Lymphocytes ($10^3$/µl)*** | 2.04 ± 1.42 | 0.56 ± 0.21 | 0.01 |
| **Platelets ($10^3$/µl)*** | 304.40 ± 125.35 | 190.97.31 | 0.09 |
| **Neutrophils ($10^3$/µl)*** | 19.59 ± 10.06 | 23.94 ± 10.23 | 0.43 |
| **Three days before death** | | | |
| **White blood cells count ($10^3$/µL)*** | 19.10 ± 8.15 | 16.42 ± 12.52 | 0.30 |
| **Hemoglobin (g/dL)*** | 11.12 ± 1.44 | 9.42 ± 1.18 | 0.52 |
| **Sodium (mmol/L)*** | 139.70 ± 6.23 | 136.56 ± 6.44 | 0.59 |
| **Potassium (mmol/L)*** | 4.79 ± 0.76 | 5.40 ± 1.37 | 0.45 |
| **Creatinine (mg/dL)*** | 2.03 ± 1.88 | 2.84 ± 2.05 | 0.95 |
| **Lymphocytes ($10^3$/µl)*** | 1.19 ± 1.07 | 1.20 ± 0.70 | 0.92 |
| **Platelets ($10^3$/µl)*** | 292.40 ± 81.96 | 188.20 93.23 | 0.05 |
| **Neutrophils ($10^3$/µl)*** | 15.84 ± 6.85 | 14.16 ± 10.81 | 0.52 |
| **Five days before death** | | | |
| **White blood cells count ($10^3$/µL)*** | 16.20 ± 6.13 | 16.36 ± 6.23 | 0.83 |
| **Hemoglobin (g/dL)*** | 10.85 ± 1.75 | 10.92 ± 1.45 | 0.91 |
| **Sodium (mmol/L)*** | 141.13 ± 5.93 | 138.06 ± 9.42 | 0.43 |
| **Potassium (mmol/L)*** | 4.51 ± 0.73 | 5.28 ± 0.58 | 0.07 |
| **Creatinine (mg/dL)*** | 2.59 ± 1.59 | 3.07 ± 1.34 | 0.51 |

| | | | |
|---|---|---|---|
| **Lymphocytes ($10^3$/µl)*** | 0.90 ± 0.87 | 0.78 ± 0.32 | 0.55 |
| **Platelets ($10^3$/µl)*** | 344.63 ± 154.60 | 205.20 ± 59.40 | 0.06 |
| **Neutrophils ($10^3$/µl)*** | 14.91 ± 4.57 | 13.15 ± 5.18 | 0.92 |

**Note:** * mean ± standard deviation; (%) percentage.

**Supplementary Table 6 - Bimodal COVID-19 Phenotype: Radiological Features.**

| CHEST XR features | Thrombotic Phenotype | Fibrotic Phenotype | P |
|---|---|---|---|
| **CHEST XR** | | | |
| **Ground-glass opacity (>50% area)** | 3 (30.0) | 4 (80.0) | 0.11 |
| **Pulmonary consolidation (%)** | 1 (10.0) | 1 (20.0) | 0.99 |
| **Pulmonary congestion (%)** | 1 (10.0) | 0 (0.0) | 0.99 |
| **Pleural effusion (%)** | 0 (0.0) | 2 (40.0) | 0.09 |
| **Interstitial opacities (>30% area)** | 5 (50.0) | 3 (60.0) | 0.99 |
| **Cardiomegaly (%)** | 6 (60.0) | 3 (60.0) | 0.99 |

**Note:** (%) percentage.

### SUPPLEMENTARY FIGURES

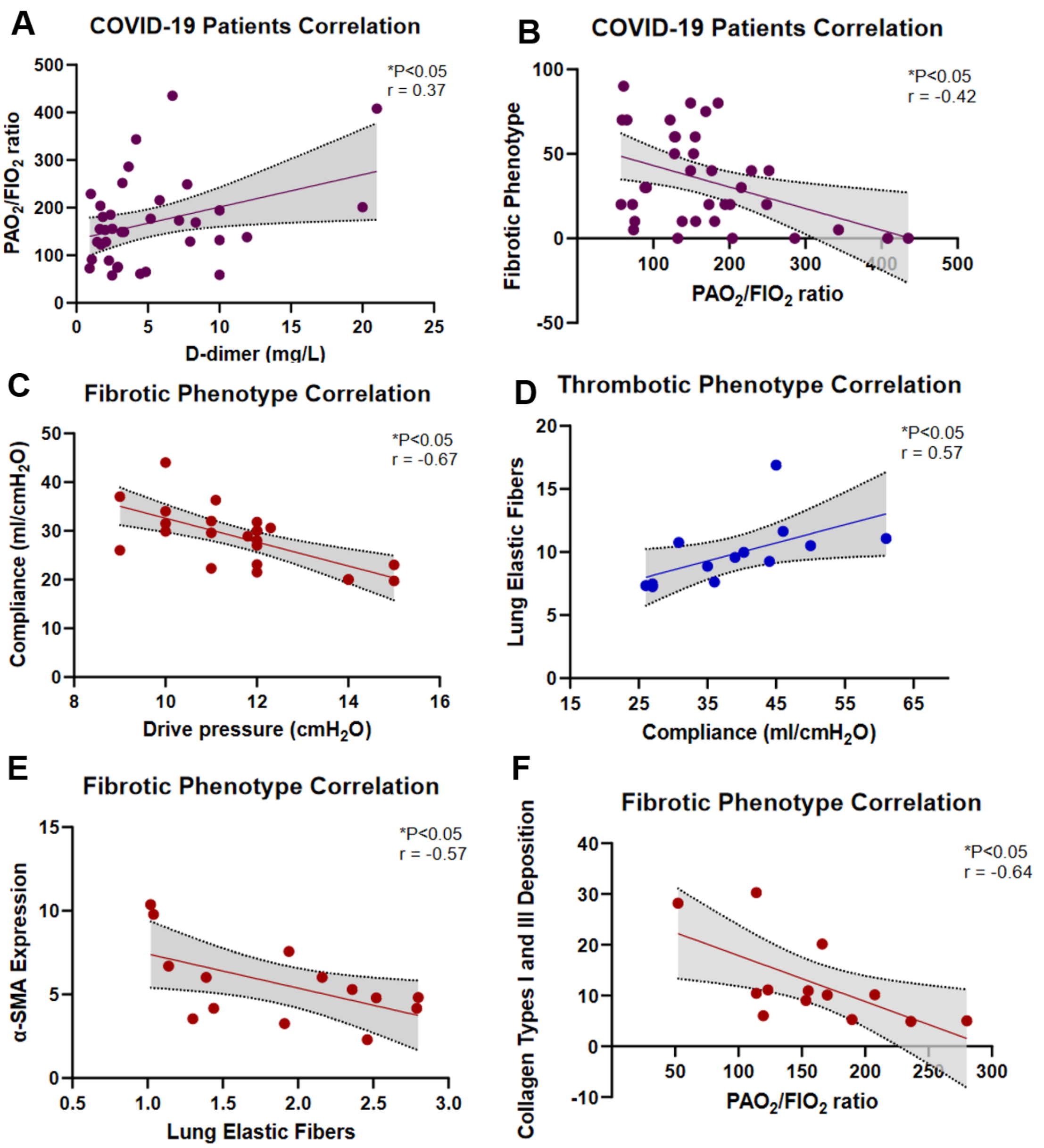


**Supplementary Figure 1 – Clinical and Pathological Data Analysis**. Following viral infection complications and subsequent mechanical ventilation, several pathophysiological processes occur and may lead to two opposite ends of lung injury manifesting as the following bimodal phenotypes: 1) Thrombotic: progressive increase in $PaO_2/FiO_2$ ratio with lung injury resolution to normal or near to normal parenchyma architecture, but prominent microthrombi formation confirmed by high D-dimer values (r=0.37; P<0.05) **(A)**; or 2) Fibrotic: progressive decline in $PaO_2/FiO_2$ ratio with lung injury resolution to interstitial fibrosis (r=-0.42; P<0.05) **(B)**. Compliance values correlated negatively with drive pressure values for fibrotic phenotype (r=-0.67; P<0.05) **(C)** and positively with lung elastic fibers, implying lung injury resolution (r=0.57; P<0.05) **(D)**. In fibrotic phenotype collagen type I and III extracellular matrix deposition by myofibroblasts activation, represented by α-SMA expression, causes stiffness of the lung parenchyma, reducing elastic capacity (r=-0.57; P<0.05) **(E)** ultimately impairing gas exchange (r=-0.64; P<0.05) **(F)**.

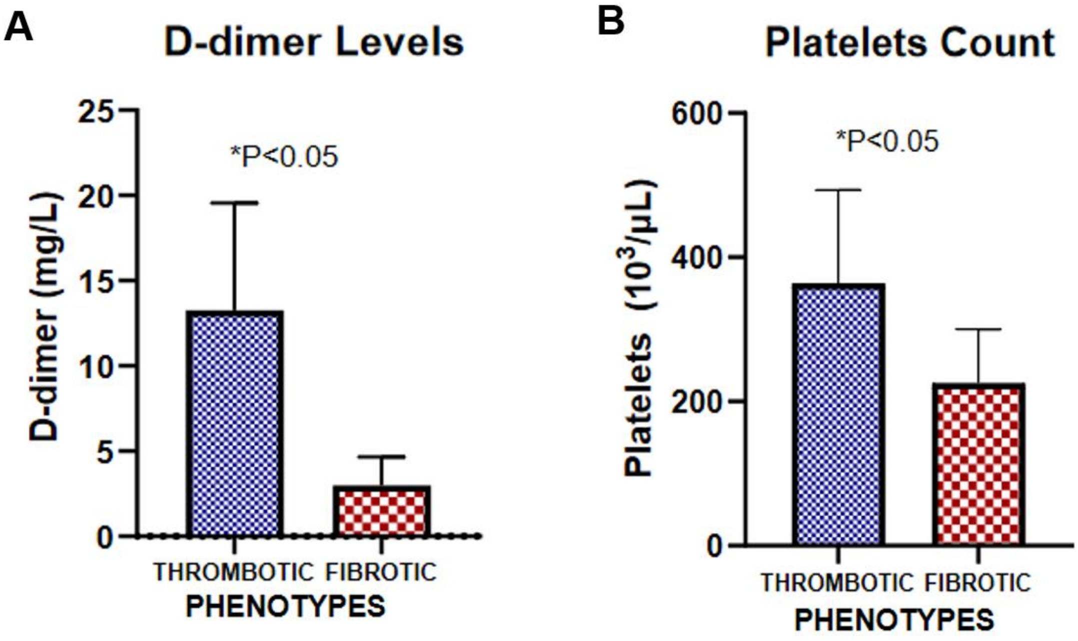


**Supplementary Figure 2 – Important statistical findings in laboratory tests.** The thrombotic phenotype showed significant increased D-dimer levels (P<0.05) **(A)** and platelets count (P<0.05) **(B)**.

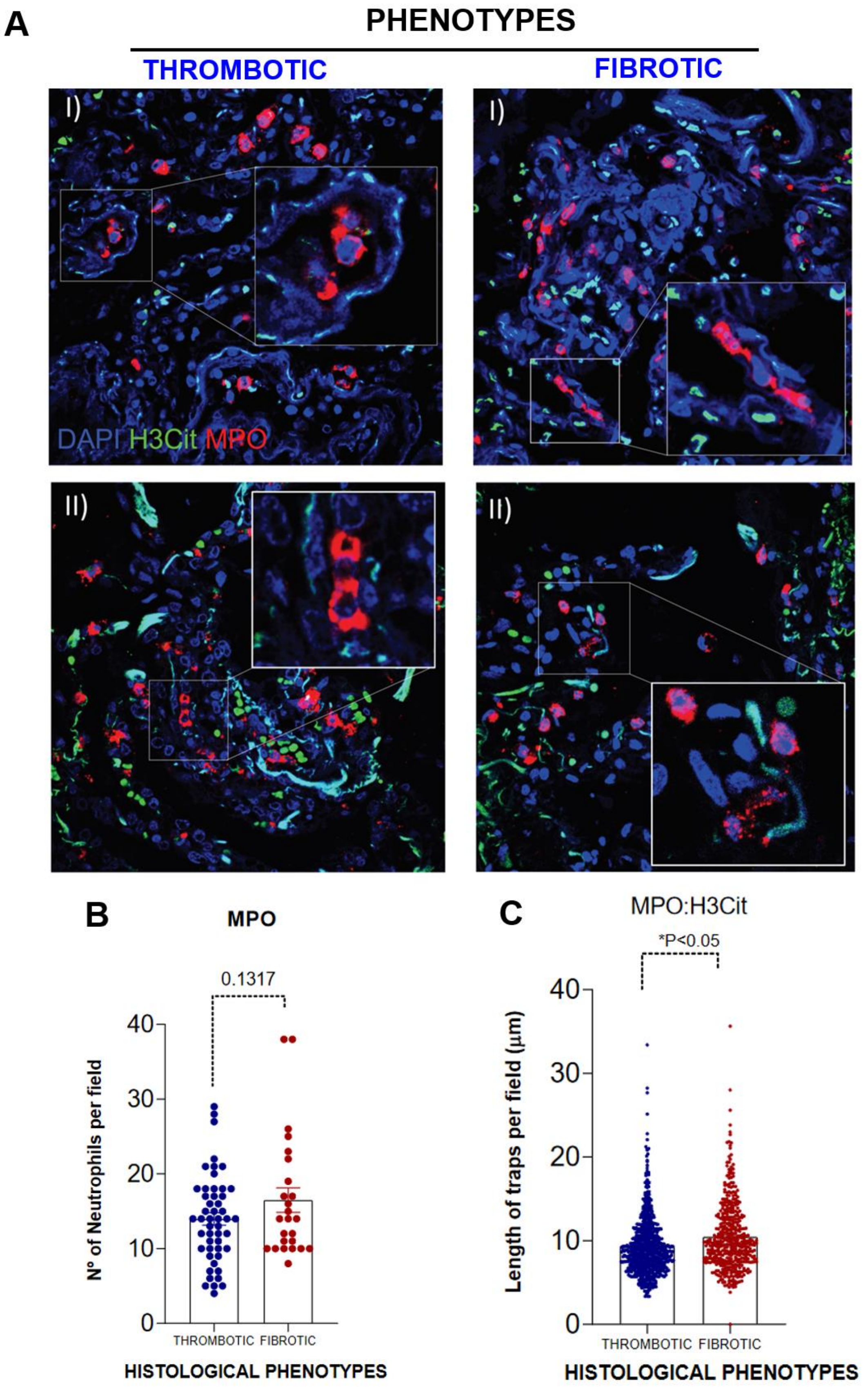
A
PHENOTYPES
THROMBOTIC
FIBROTIC
I)
II)
DAPI H3Cit MPO
B
MPO
0.1317
Nº of Neutrophils per field
0
10
20
30
40
THROMBOTIC FIBROTIC
HISTOLOGICAL PHENOTYPES
C
MPO:H3Cit
*P<0.05
Length of traps per field (μm)
0
10
20
30
40
THROMBOTIC FIBROTIC
HISTOLOGICAL PHENOTYPES

**Supplementary Figure 3 – Neutrophil-derived extracellular traps in fibrotic phenotype.** Immunofluorescence identified neutrophils with no significant quantitative difference between fibrotic and thrombotic phenotypes **(A, B)**. However, NETs were significantly higher in lung parenchyma from fibrotic phenotype subjects. ($P<0.05$) **(C)**.
